## Supplemental Figures S1-S15 for "Genetic control of fetal placental genomics contributes to development of health and disease"

**Figure S1: SNP heritability of 40 traits.** Estimates of SNP heritability with 95% confidence interval (X-axis), grouped and colored by trait category (Y-axis).

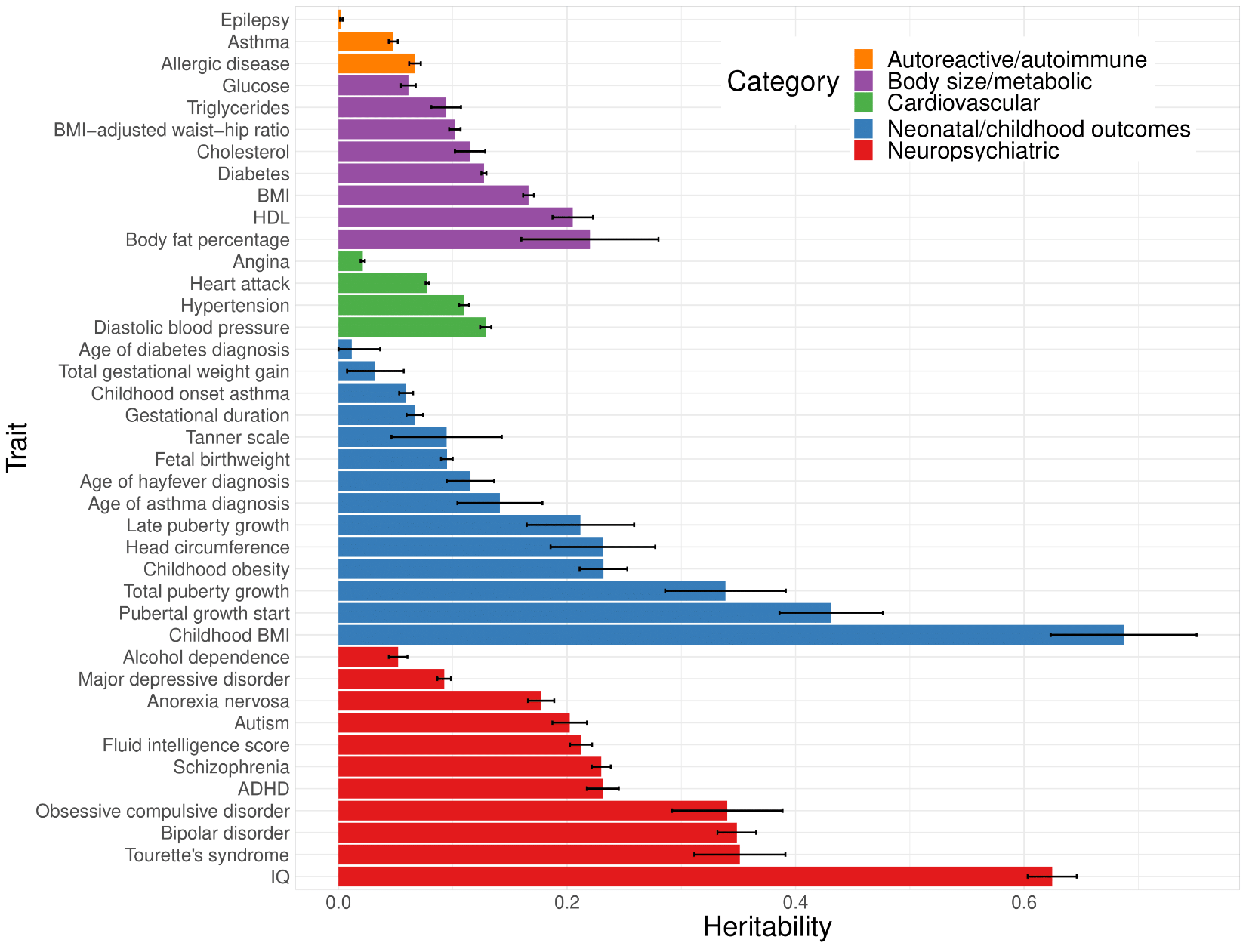

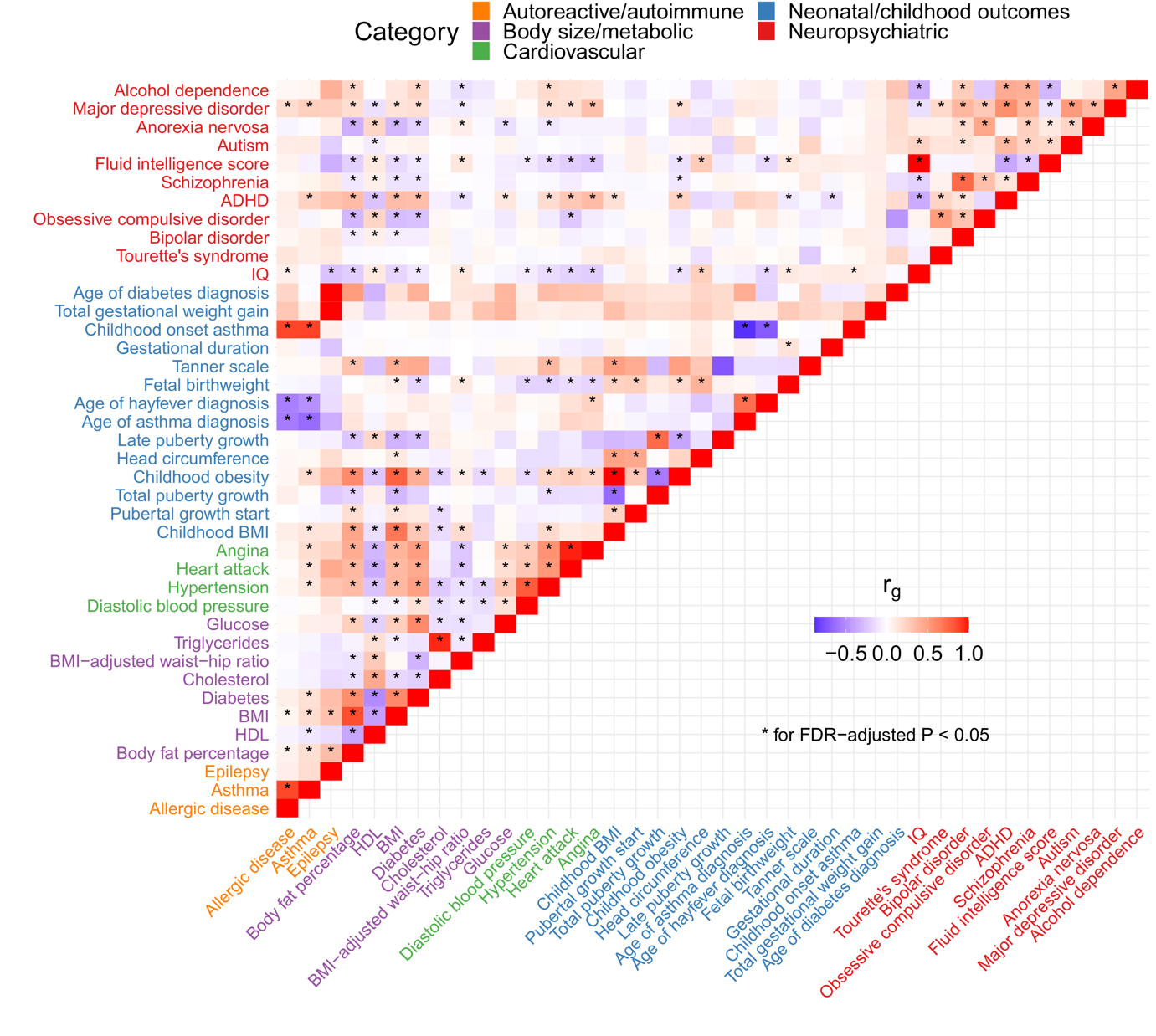
**Figure S2: SNP-based genetic correlation between 40 traits.** Heatmap of estimates of SNP-based genetic correlated between traits, grouped and colored by trait category. Correlations are marked with an asterisk are significantly non-zero with FDR-adjusted $P < 0.05$.

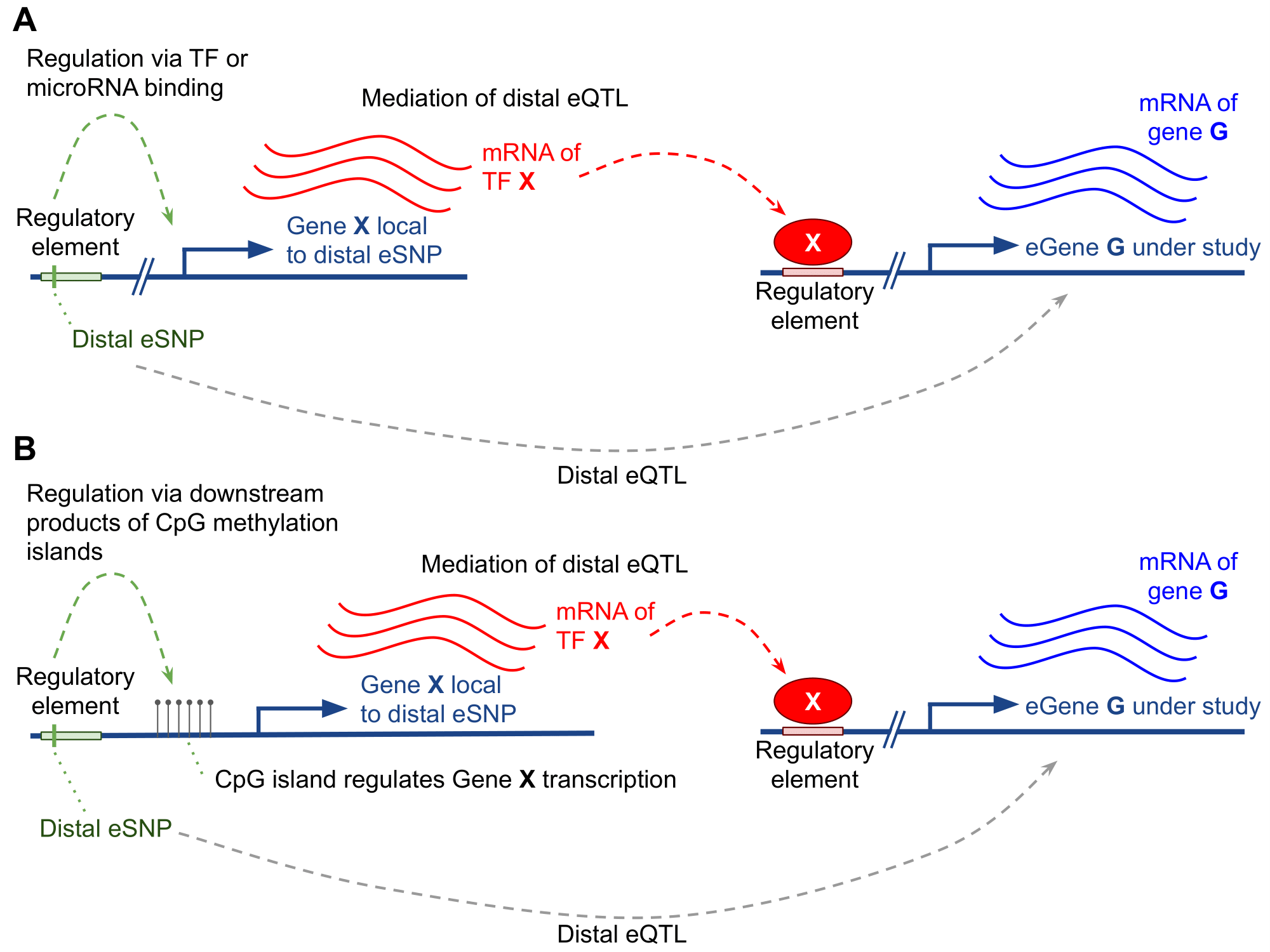

**Figure S3**: **Example of a biological mechanism MOSTWAS leverages in its predictive models**. Here, assume a SNP (in green) within a regulatory element affects the transcription of gene X (A) or the hyper- or hypomethylation of a CpG island upstream of gene X (B) that codes for a transcription factor or a microRNA hairpin. Transcription factor or microRNA X then binds to a distal regulatory region and affects the transcription of gene G. The association between the expression of gene X and gene G is leveraged in the first step of MeTWAS. A distal-eQTL association is also conferred between this distal-SNP and the eGene G, which is leveraged in the DePMA training process.

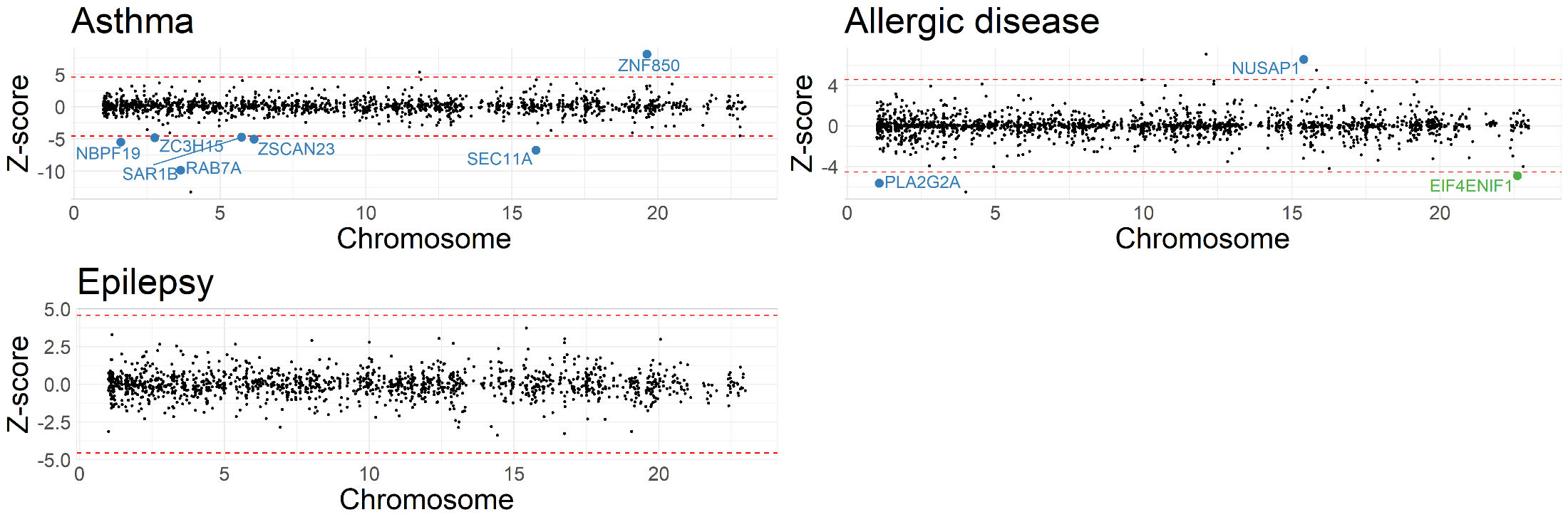
**Figure S4: TWAS Miami plots for autoimmune/autoreactive disorders**. Weighted Z-scores for TWAS associations (Y-axis) over genomic location of genes (X-axis). Red lines show Z-scores corresponding to P < $2.5\times{10}^{-6}$. Genes labelled have P < $2.5\times{10}^{-6}$, nominal permutation P <0.05, and genes in green showed Benjamini-Hochberg FDR-adjusted P < 0.05 for the distal-SNPs added-last test. All traits represented here are measured in adults*.*

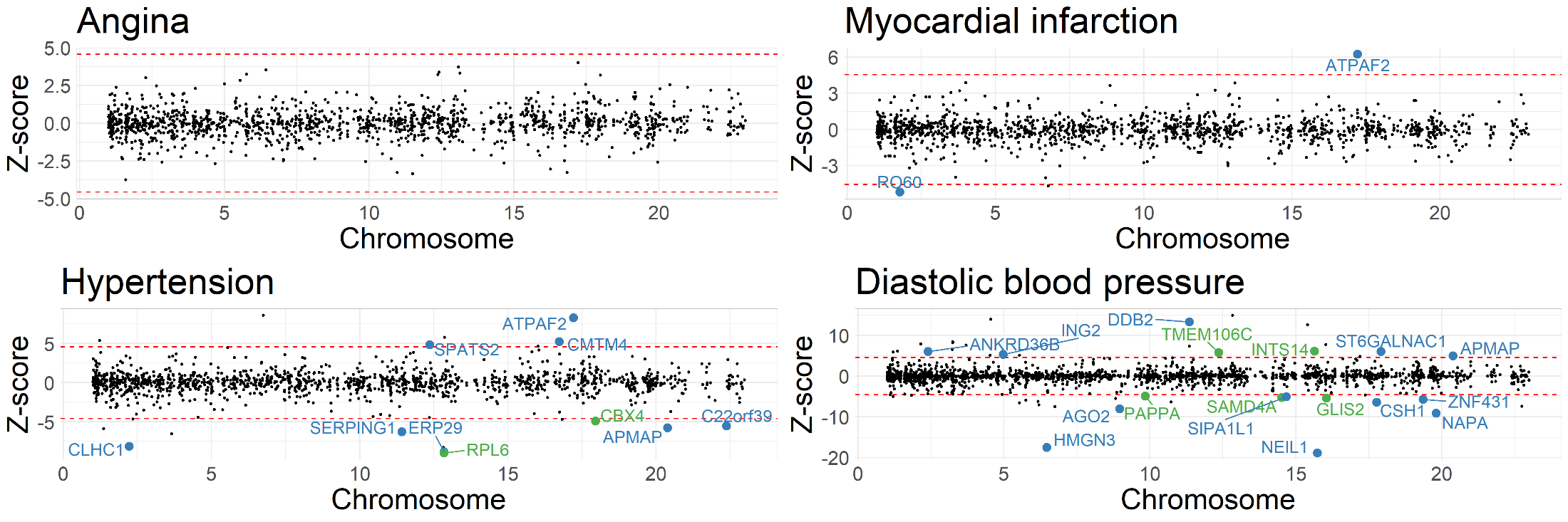
**Figure S5: TWAS Miami plots for cardiovascular disorders**. Weighted Z-scores for TWAS associations (Y-axis) over genomic location of genes (X-axis). Red lines show Z-scores corresponding to P < $2.5\times{10}^{-6}$. Genes labelled have P < $2.5\times{10}^{-6}$, nominal permutation P <0.05, and genes in green showed Benjamini-Hochberg FDR-adjusted P < 0.05 for the distal-SNPs added-last test. All traits represented here are measured in adults*.*

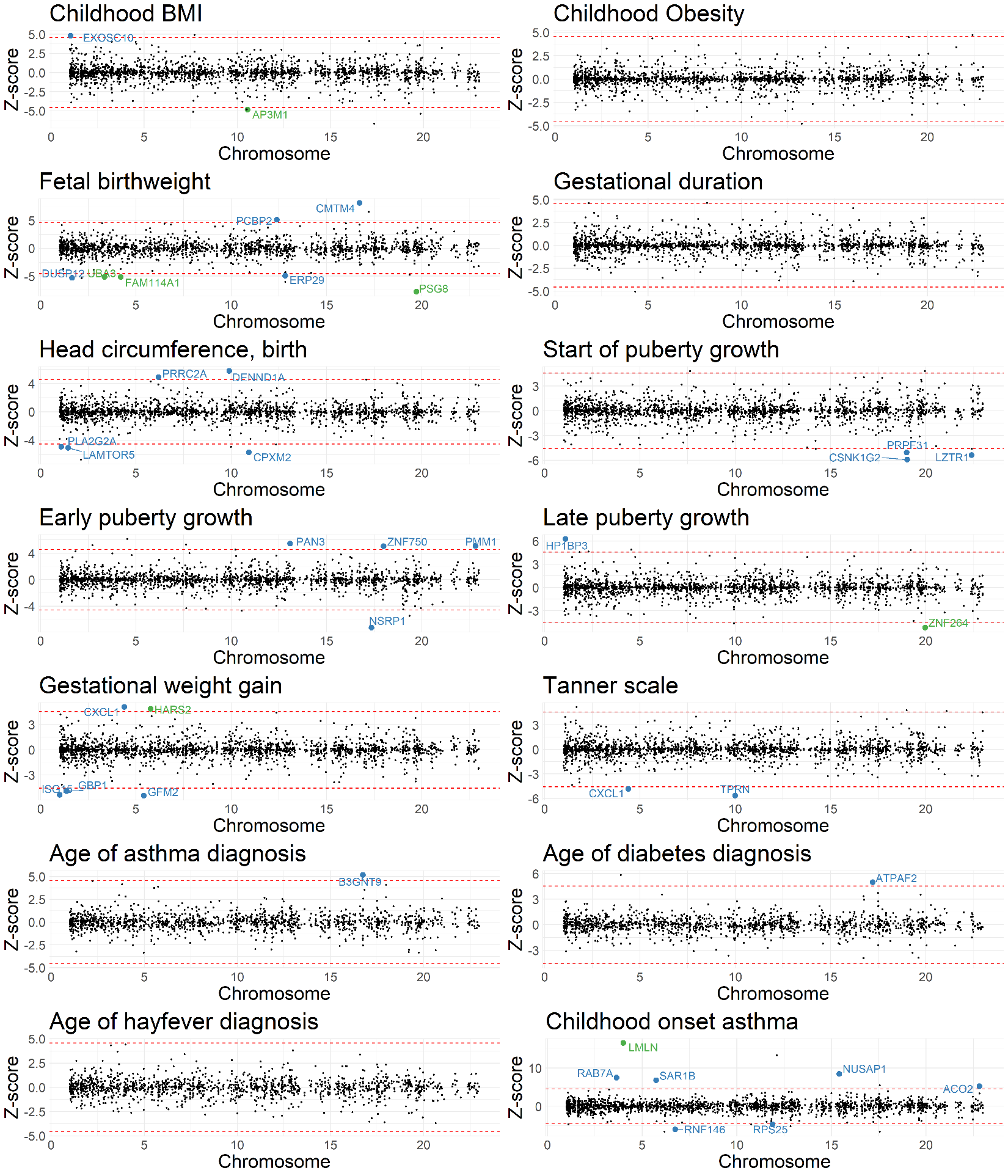

**Figure S6: TWAS Miami plots for neonatal/childhood outcomes**. Weighted Z-scores for TWAS associations (Y-axis) over genomic location of genes (X-axis). Red lines show Z-scores corresponding to P < $2.5\times{10}^{-6}$. Genes labelled have P < $2.5\times{10}^{-6}$, nominal permutation P < 0.05, and genes in green showed Benjamini-Hochberg FDR-adjusted P < 0.05 for the distal-SNPs added-last test. All traits represented here are measured in infants or children.

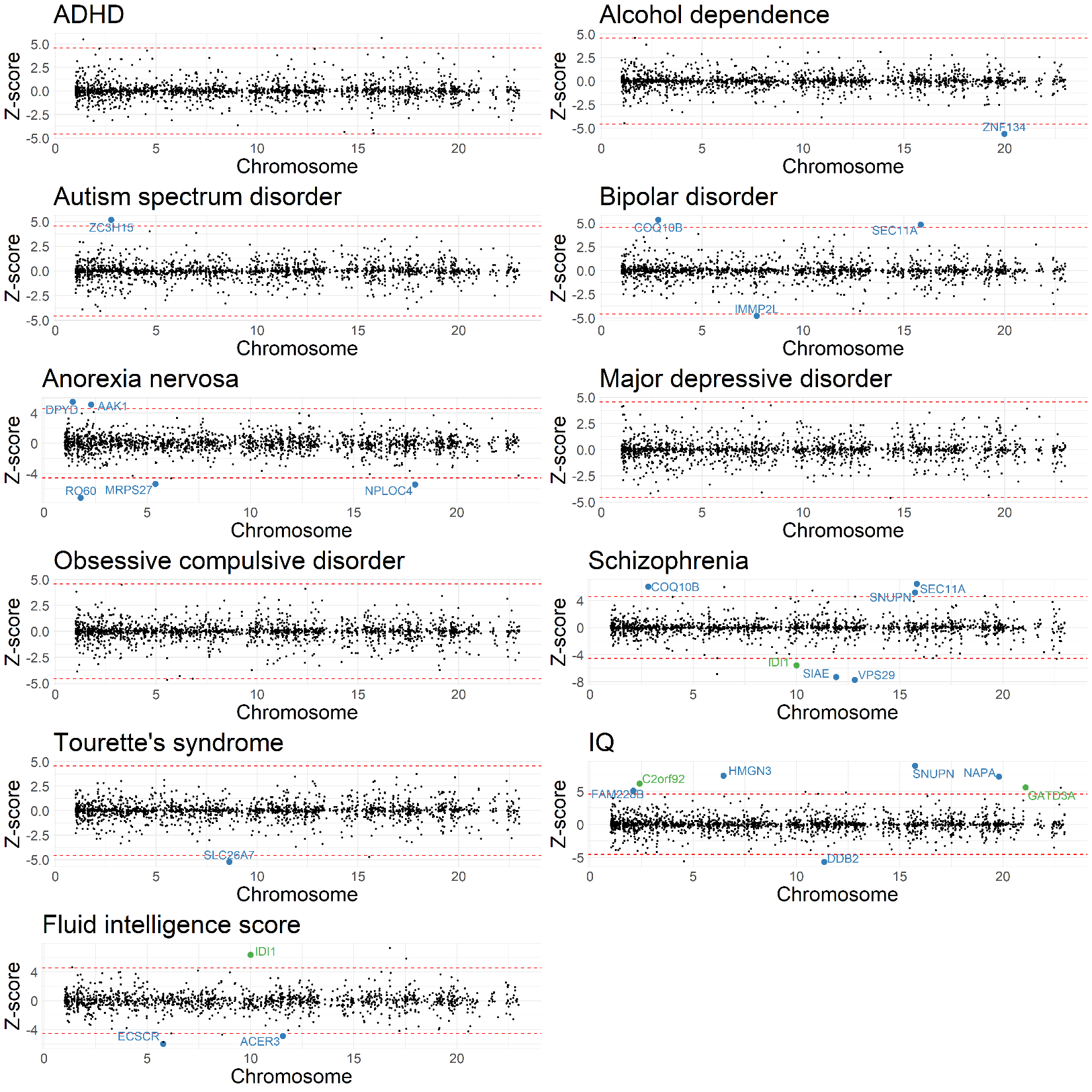

**Figure S7: TWAS Miami plots for neuropsychiatric outcomes**. Weighted Z-scores for TWAS associations (Y-axis) over genomic location of genes (X-axis). Red lines show Z-scores corresponding to P < $2.5\times{10}^{-6}$. Genes labelled have P < $2.5\times{10}^{-6}$, nominal permutation P < 0.05, and genes in green showed Benjamini-Hochberg FDR-adjusted P < 0.05 for the distal-SNPs added-last test.

All traits represented here are measured in adults*.*

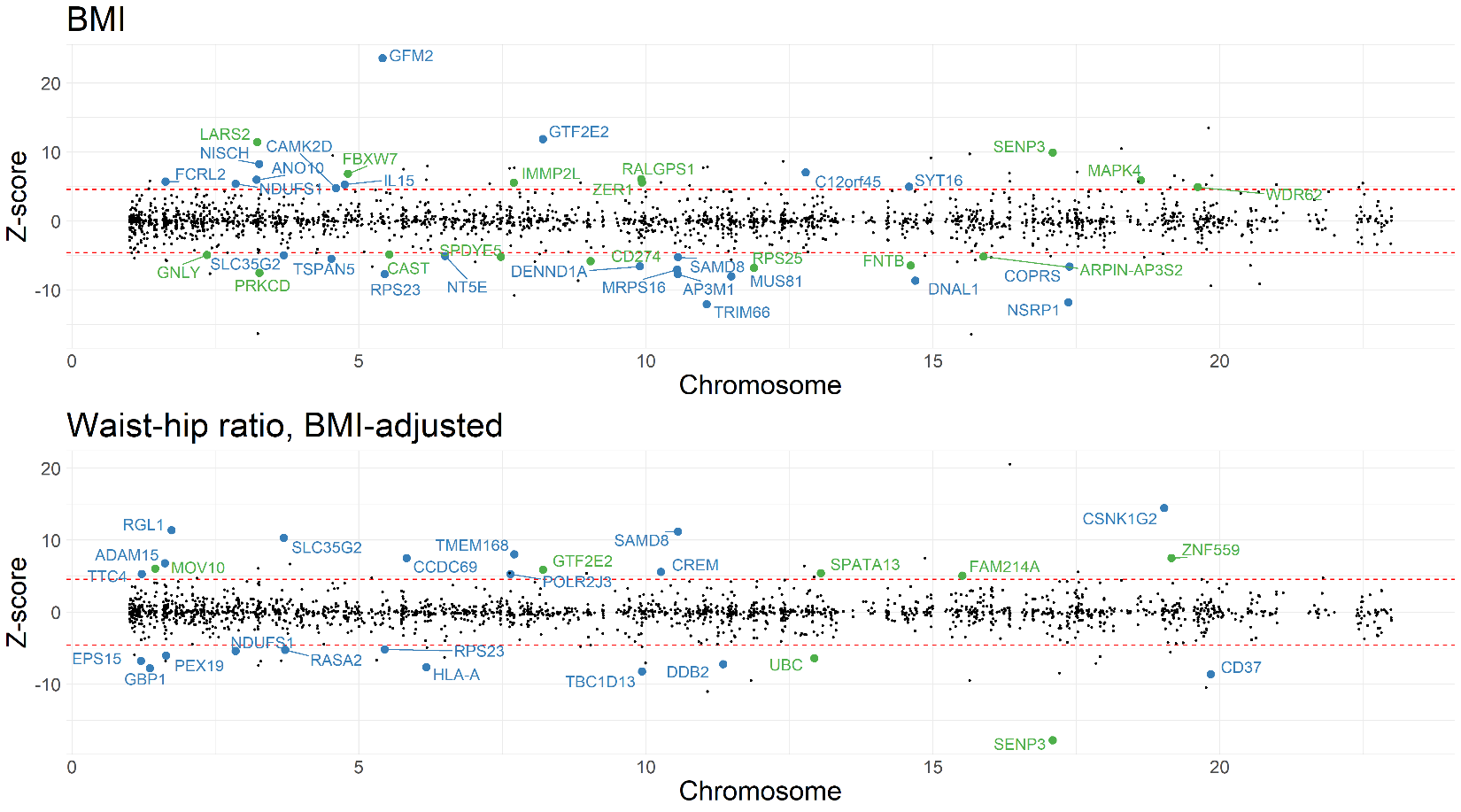
**Figure S8: TWAS Miami plots for BMI and BMI-adjusted waist-hip ratio**. Weighted Z-scores for TWAS associations (Y-axis) over genomic location of genes (X-axis). Red lines show Z-scores corresponding to P < $2.5\times{10}^{-6}$. Genes labelled have P < $2.5\times{10}^{-6}$, nominal permutation P <0.05, and genes in green showed Benjamini-Hochberg FDR-adjusted P < 0.05 for the distal-SNPs added-last test. All traits represented here are measured in adults*.*

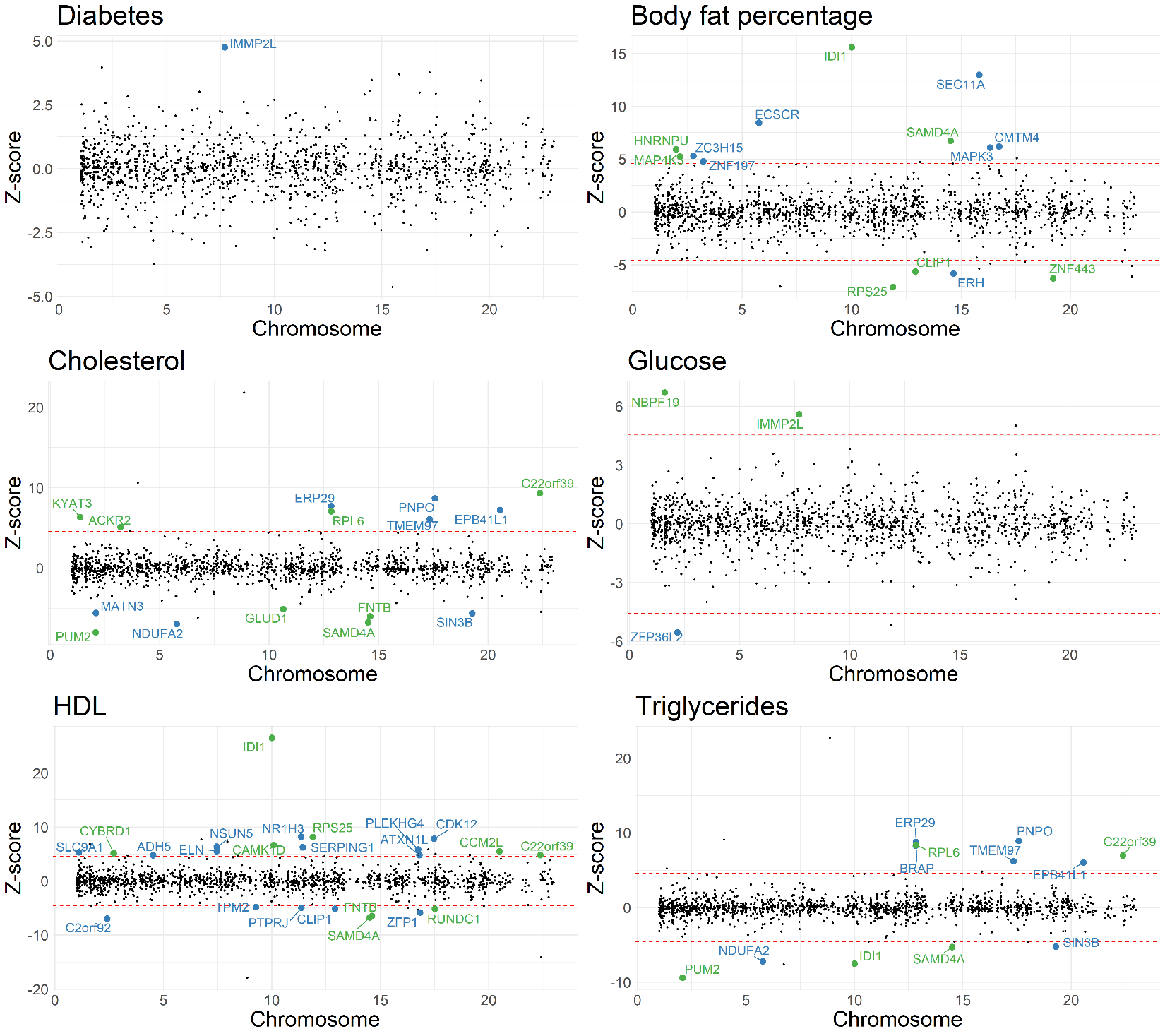
**Figure S9: TWAS Miami plots for body size/metabolic traits, excluding BMI and BMI-adjusted waist-hip ratio**. Weighted Z-scores for TWAS associations (Y-axis) over genomic location of genes (X-axis). Red lines show Z-scores corresponding to P < $2.5\times{10}^{-6}$. Genes labelled have P < $2.5\times{10}^{-6}$, nominal permutation P <0.05, and genes in green showed Benjamini-Hochberg FDR-adjusted P < 0.05 for the distal-SNPs added-last test. All traits represented here are measured in adults.

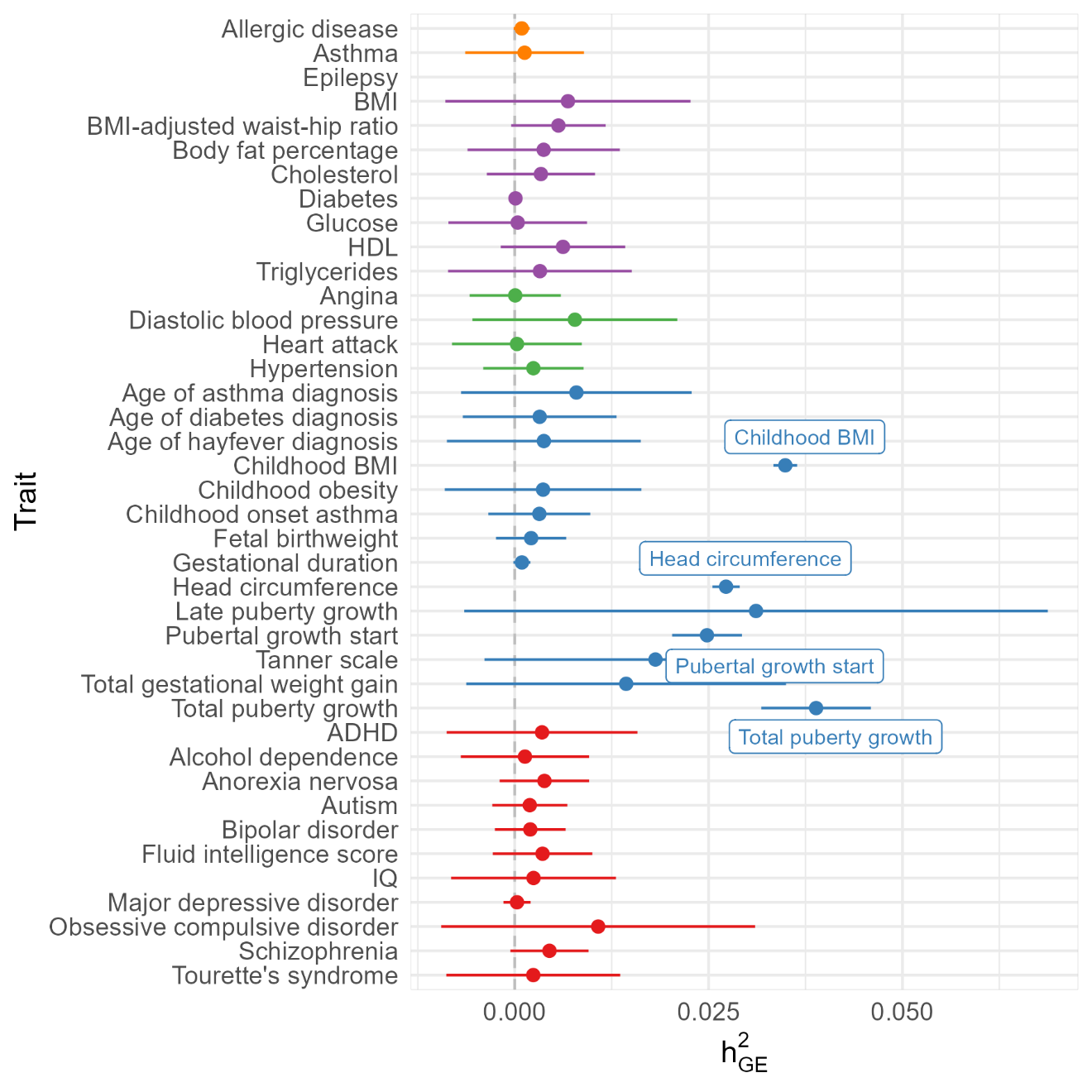

**Figure S10:** Placental expression-mediated genetic heritability of traits. Caterpillar plot of placental expression-meidated genetic heritability of traits, colored by trait category. Wald-type 95% confidence intervals are provided for reference. Trait is labelled if the confidence interval does not intersect the null of $h_{GE}^{2}=0$.

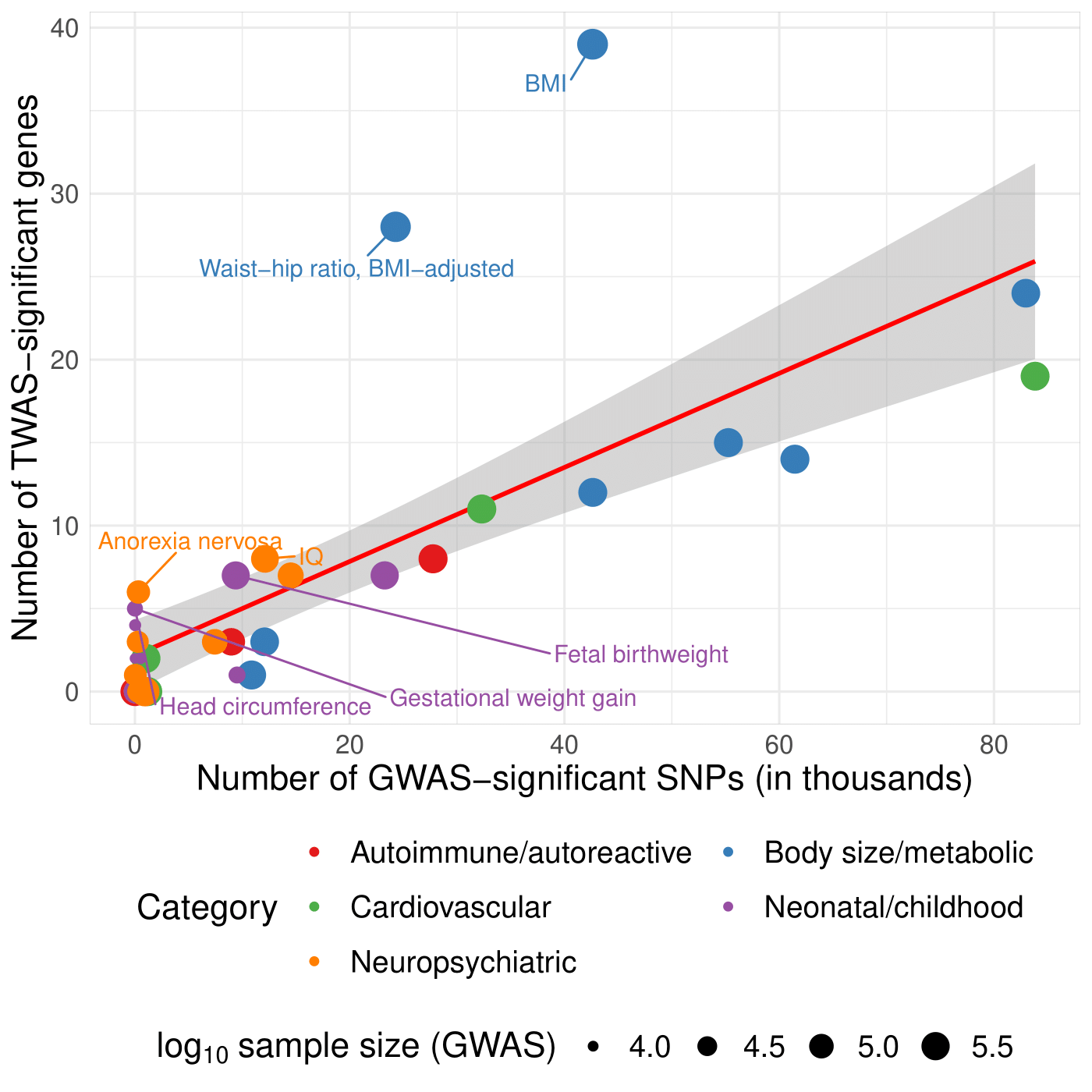

**Figure S11: Comparison of GWAS and TWAS results across all 40 traits**. Scatterplot of number of TWAS-significant genes (Y-axis) and number of GWAS-significant SNPs (X-axis) across all 40 traits, colored by category of the trait. The size of the point shows the log_10_ sample size of the GWAS. The red line and gray band provide a regression line and 95% confidence band for the fitted values. Points are labelled if the point falls outside the confidence band.

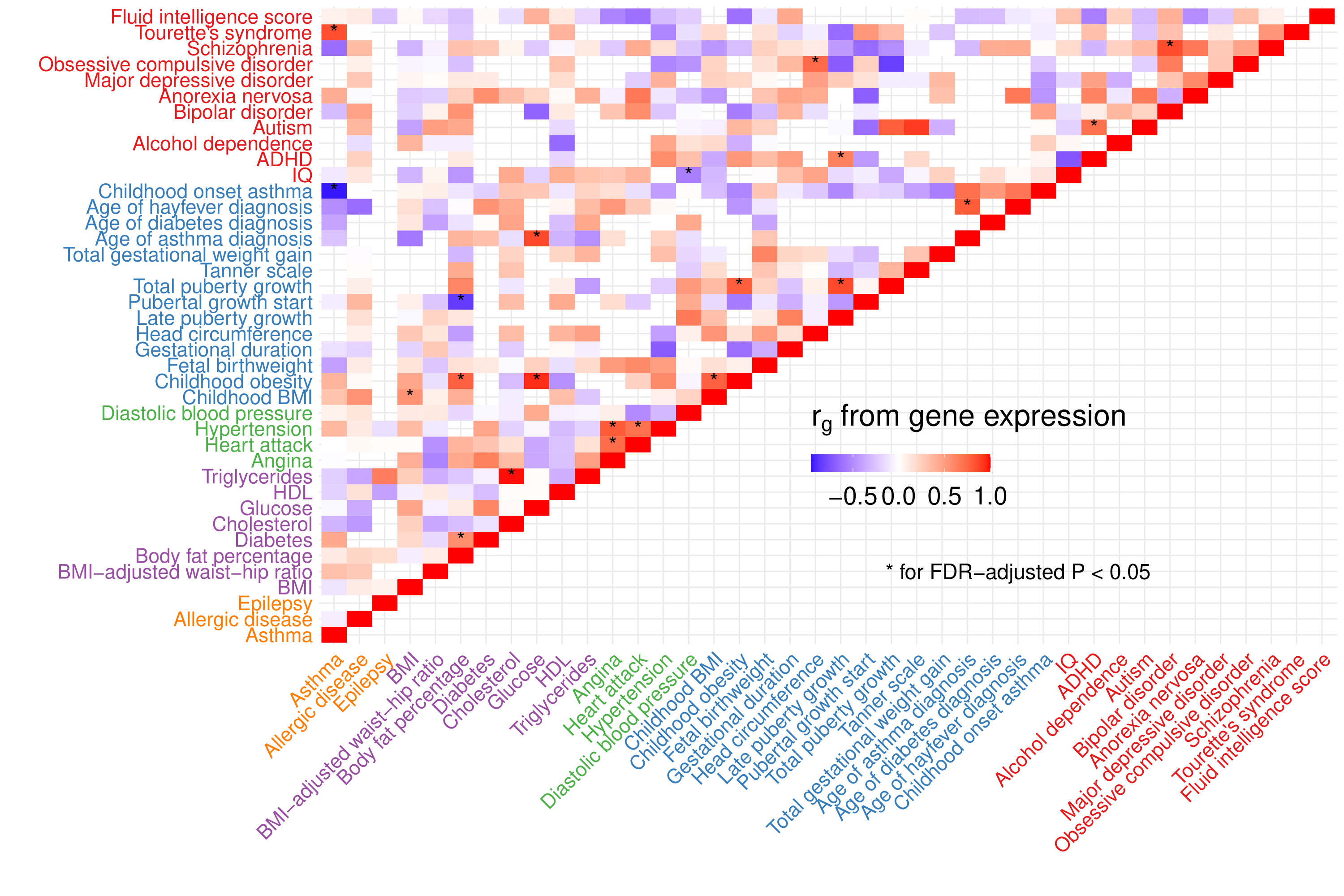
**Figure S12: Heatmap of genetic correlations on the heritable gene expression level between 40 traits considered in TWAS analysis**. Genetic correlations between traits at the level of the predicted expression of heritable genes. Correlations at FDR-adjusted P <0.05 are marked with an asterisk. Autoimmune/autoreactive traits are colored in yellow, body size/metabolic in purple, cardiovascular in green, neonatal/childhood outcomes in blue, and neuropsychiatric in red.

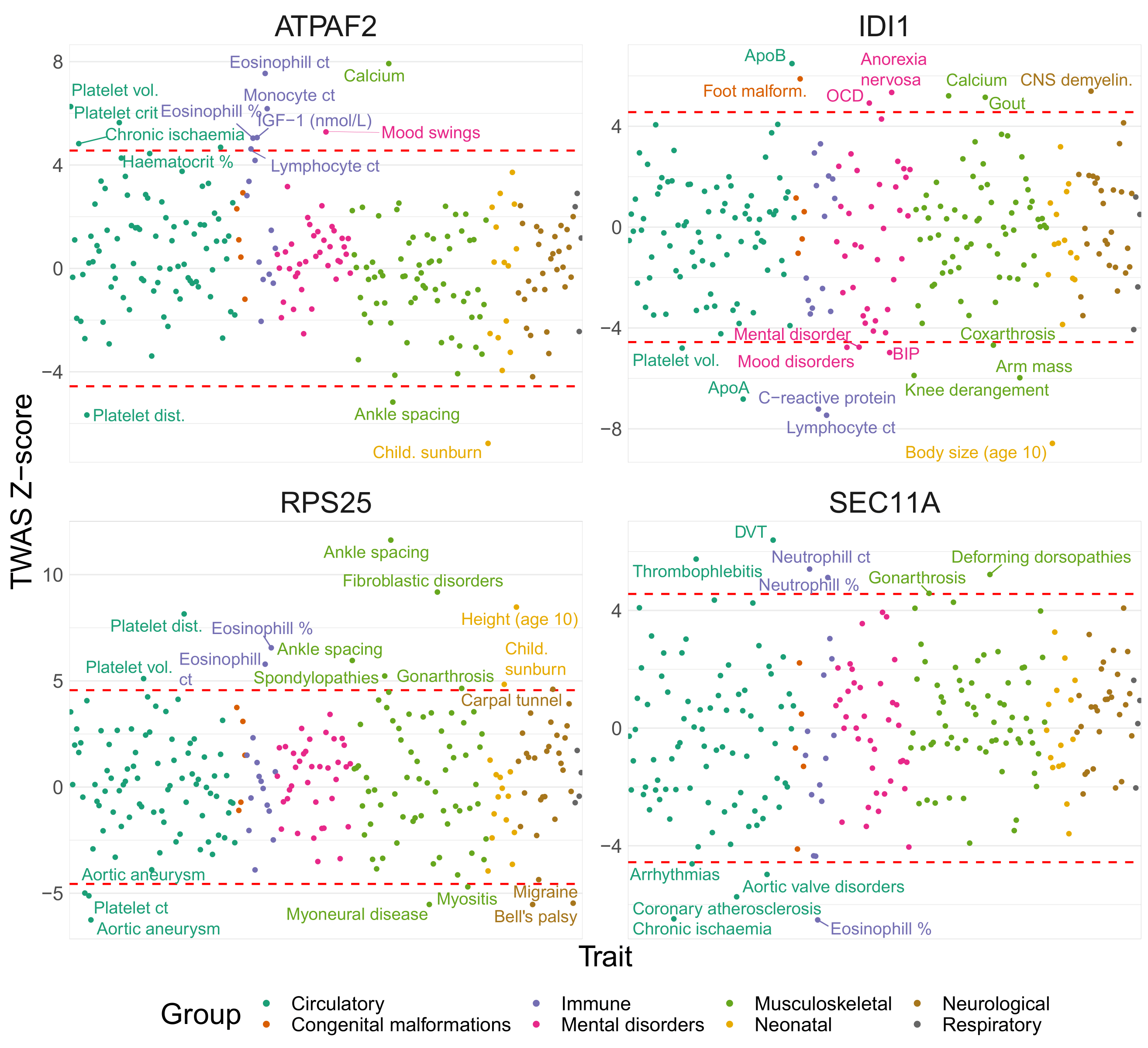

**Figure S13: Miami plot of representative phenome-wide scans of GTAs in UKBB**. Weighted burden Z-score (Y-axis) of GTA across all traits (X-axis), grouped and colored by ICD code block.

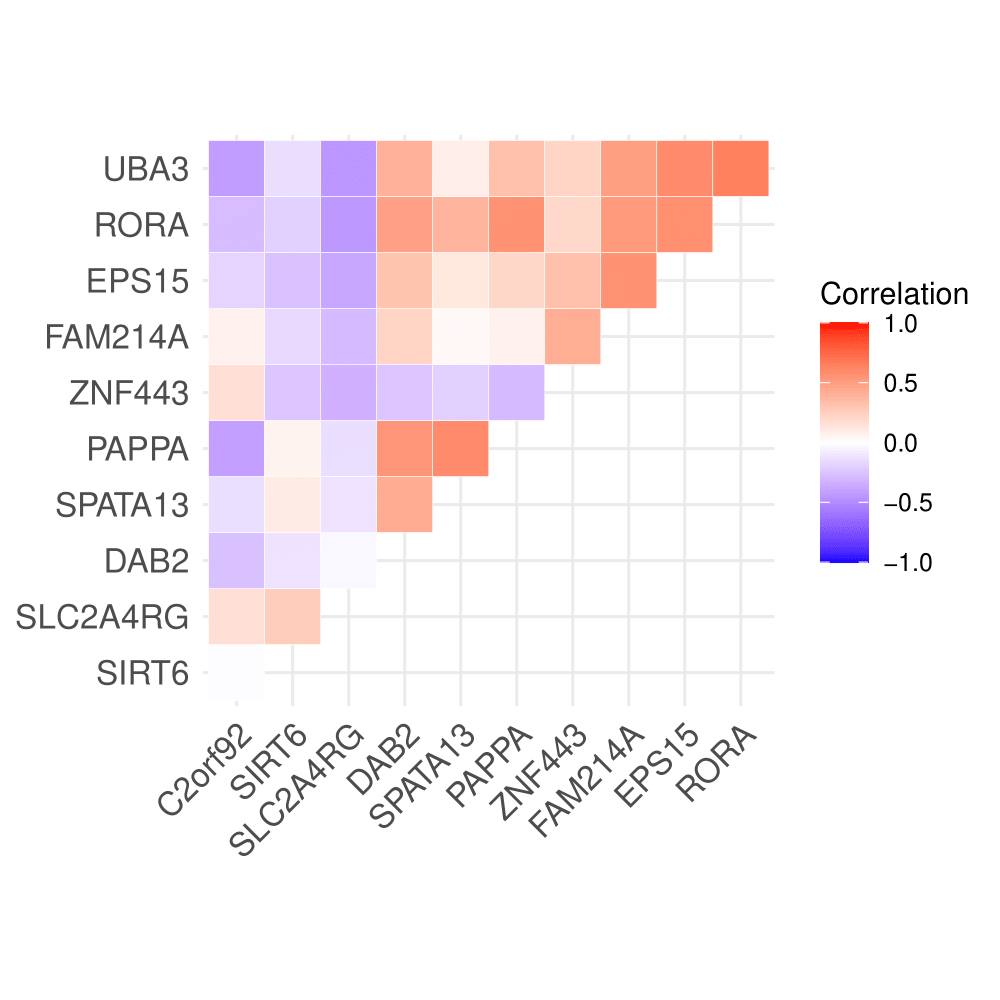

**Figure S14: Heatmap of correlations between select transcription factor and TWAS-identified genes in RICHS**. Correlations between the RICHS expression of RPs (Y-axis) and associated TWAS genes identified by MOSTWAS in ELGAN (X-axis).

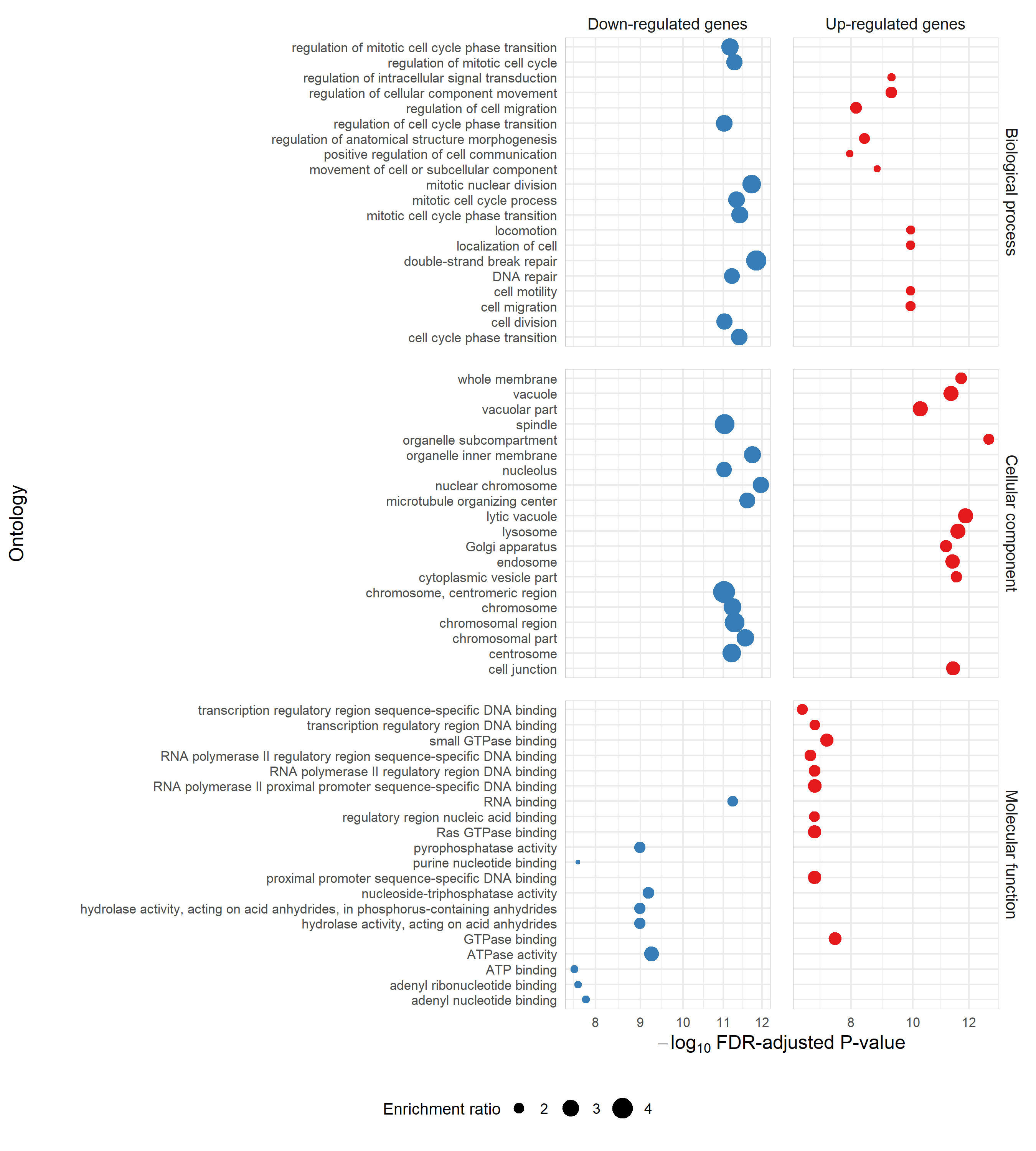
**Figure S15: Over-representation enrichments of differentially expressed genes in *EPS15* knockdown.** Enrichment plot of over-representation of biological process, cellular component, and molecular function ontologies (Y-axis) with -log_10_ FDR-adjusted P-value (X-axis). The size of the point gives the relative enrichment ratio for the given pathway.
